# Custommune: a web tool to design personalized and population-targeted vaccine epitopes

**DOI:** 10.1101/2020.04.25.20079426

**Authors:** Mohammad Tarek, Mahmoud Elhefnawi, Juliana Terzi Maricato, Ricardo Sobhie Diaz, Iart Luca Shytaj, Andrea Savarino

**Affiliations:** Bioinformatics Department, Armed Forces College of Medicine (AFCM), Cairo, Egypt; Biomedical Informatics and Chemo-Informatics Group, Centre of Excellence for medical research, Informatics and Systems Department, National Research Centre, Cairo, Egypt; Federal University of Sao Paulo, Infectious Diseases Department, São Paulo, Brazil; Department of Infectious Diseases, Integrative Virology, University Hospital Heidelberg, Heidelberg, Germany; Department of Infectious Diseases, Italian Institute of Health, Rome, Italy

## Abstract

Computational prediction of immunogenic epitopes is a promising platform for therapeutic and preventive vaccine design. A potential target for this strategy is human immunodeficiency virus (HIV-1), for which, despite decades of efforts, no vaccine is available. In particular, a therapeutic vaccine devised to eliminate infected cells would represent a key component of cure strategies. HIV peptides designed based on individual viro-immunological data from people living with HIV/AIDS have recently shown able to induce post-therapy viral set point abatement. However, the reproducibility and scalability of this method is curtailed by the errors and arbitrariness associated with manual peptide design as well as by the time-consuming process.

We herein introduce Custommune, a user-friendly web tool to design personalized and population-targeted vaccines. When applied to HIV-1, Custommune predicted personalized epitopes using patient specific Human Leukocyte Antigen (HLA) alleles and viral sequences, as well as the expected HLA-peptide binding strength and potential immune escape mutations. Of note, Custommune predictions compared favorably with manually designed peptides administered in a recent phase II clinical trial (NCT02961829).

Furthermore, we utilized Custommune to design preventive vaccines targeted for populations highly affected by COVID-19. The results allowed the identification of peptides tailored for each population and predicted to elicit both CD8^+^ T-cell immunity and neutralizing antibodies against structurally conserved epitopes of severe acute respiratory syndrome coronavirus 2 (SARS-CoV-2).

Overall, our data describe a new tool for rapid development of personalized or population-based immunotherapy against chronic and acute viral infections.

## Introduction

The rapid development of automated platforms for data generation and analysis are increasingly making precision medicine a concrete option for several diseases. Due to its potential for high selectivity and efficacy, immunotherapy is an optimal choice for the design of personalized therapeutic interventions^1^. While most efforts in this direction have focused on cancer^1–3^, viral infections can be a relevant application as well, particularly chronic infections characterized by extensive genetic diversity, in part due to in-host viral evolution.

Human immunodeficiency virus (HIV-1) is case in point, as the large number of circulating strains and its high replicative mutation rates have hampered the development of effective vaccines, both preventive and therapeutic^4,5^. Several lines of evidence highlight the relevance of immune control in HIV-1 infection. Spontaneous long-term control of HIV-1 replication can be accompanied by the presence of broadly neutralizing antibodies^6,7^ or, more frequently, effective cell-mediated immune responses^8^. Moreover, protective Class I HLA alleles have been identified both in people living with HIV/AIDS (PLWHA) and macaques infected with the HIV homolog simian immunodeficiency virus (SIV)^9–14^. In line with this, temporary depletion of CD8^+^ T-cells is associated with a rapid viral load increase, while their replenishment can revert this effect^15–18^.

A therapeutic vaccine based on cell-mediated immunity might offer the advantage of decreasing the number of infected cells. On the one hand, HIV-1 latently infected cells, which constitute the main barrier to a cure^19–21^, are not targeted by antiretroviral drugs or CD8^+^ T-cells^22^. On the other hand, effective cell-mediated immune responses could preserve drug-free control of the infection by keeping viral load low/undetectable and by eliminating the infected cells undergoing spontaneous HIV-1 reactivation from latency. Such therapeutic vaccines could also be combined with strategies aimed at purging the HIV-1 latent reservoirs by inducing pharmacologic reactivation of latently infected cells^23^.

The strong correlation between the host’s genetic background and immune-mediated control of the infection suggests that effective immunity is mainly directed against a subset of HIV-1 epitopes. Consistently, several studies have shown that cell-mediated immune responses against the HIV-1 Gag protein correlate with lower viral loads in PLWHA and with post-therapy control of the infection in macaques^18,24–27^. The peculiar efficacy of anti-Gag immunity might be partially explained by the higher fitness cost associated with mutations in this viral protein^28^. In particular, specific regions of Gag, which are essential for HIV-1 packaging and assembly, are structurally and evolutionarily conserved, displaying low Shannon entropy both in humans and primate lentiviruses^29^. However, it is noteworthy that low diversity is not sufficient *per se* to induce viral load control, as vaccine approaches designed exclusively by selecting epitopes based on their evolutionary conservation have shown only modest effects^30,31^.

A recent phase II clinical trial (NCT02961829) has attempted to induce anti-Gag immunity against conserved epitopes using a personalized approach based on patient HLA sequences^32^. Although the study enrolled only a small number of PLWHA and tested multiple interventions, preliminary results suggest that therapeutic vaccination with autologous dendritic cells pulsed with individually designed peptides decreased the viral set point in some patients during analytical treatment interruption (ATI)^32^.

In the present work we describe and test a new automated, user-friendly web-based tool to design personalized peptides for vaccination. The tool, named Custommune, was principally interrogated to develop therapeutic vaccine candidates for HIV-1. To this aim, by intersecting input data from patient-specific viral sequences and HLA alleles, Custommune provides an output of epitopes of desired length filtered for their predicted specificity, immunogenicity and mutation potential. Of note, in our simulations, Custommune performance was superior to that of manual vaccine design (applied in clinical trial NCT02961829) in terms of prediction of clinical response.

One advantage of Custommune over traditional vaccine design techniques is the ability to quickly adapt the tool for different targets and strategies. In this regard, we applied Custommune to the novel pandemic COVID19, caused by severe acute respiratory syndrome coronavirus 2 (SARS-CoV-2)^33^. Due to the acute manifestations of the disease, the design of a population-targeted preventive vaccine was chosen as a more practical approach as compared to a personalized therapeutic vaccine. Moreover, to broaden the expected coverage and increase the likelihood of achieving herd immunity^34^, a strategy able to potentially evoke both neutralizing antibodies and cell-mediated immunity was preferred. Using input regions of SARS-CoV-2 identified as viable targets, Custommune was able to design vaccine candidates specific for regionally prevalent HLA genotypes. In addition, Custommune selected those HLA Class II restricted epitopes that could induce neutralizing antibodies and thus provide a two-layered protection against the infection.

Taken as a whole, our results show the potential of the Custommune algorithm to quickly design personalized or population-specific peptides for preventive and therapeutic vaccination. Due to its intuitive and scalable approach, Custommune might provide an effective tool for rapid vaccine development against chronic and acute conditions.

## Results

### Custommune pipeline for prediction of candidate vaccine peptides

The Custommune web tool (available at: http://www.custommune.com) was written in Python (http://www.python.org) using the Django framework (https://www.djangoproject.com) and provides the user with an easy online interface for accessing and downloading prediction datasets without any coding knowledge requirements. The tool utilizes a pipeline (Figure 1) to design epitopes for preventive and therapeutic vaccines.

**Figure 1.**
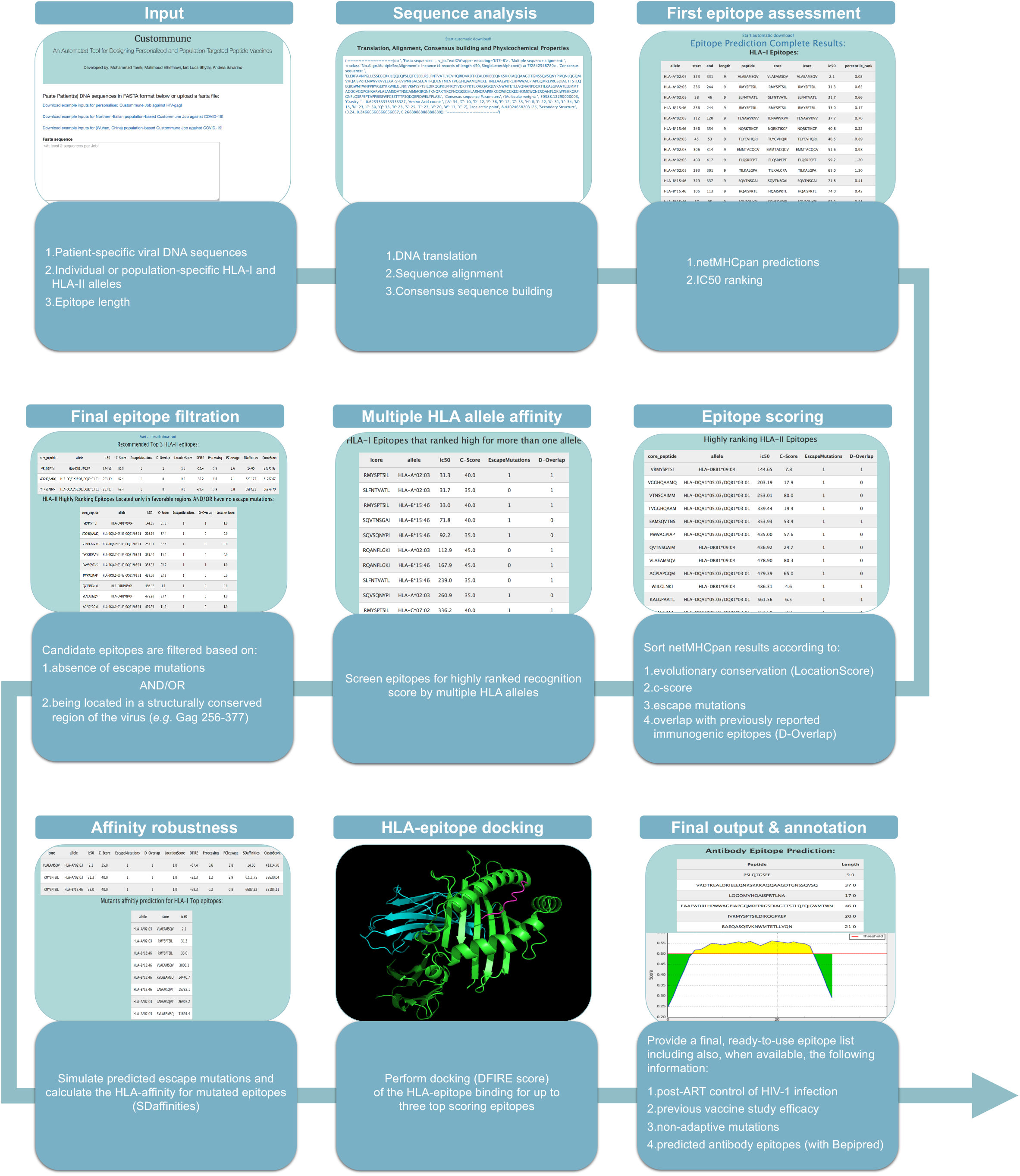
Illustrated workflow of Custommune epitope prediction pipeline. (*Input*) the Custommune pipeline starts by validating user inputs for sequences, alleles and desired epitope length. (*Sequence analysis*) input sequences are then translated to build an alignment of amino acid sequences from which a consensus sequence is generated and used for further epitope prediction. (*First epitope assessment*) using the netMHCpan 4.0 algorithm^35^, Custommune initially ranks epitope predictions based on their IC50 values. (*Epitope scoring*) additional scoring layers are then applied by Custommune based on: location of the epitope (by assigning a LocationScore to epitopes located in an evolutionary conserved region); evolutionary conservation of the epitope residues (C-Score) assessed by using an internal sequence database (Supplementary File 1) or the Basic Local Alignment Search Tool (BLAST; https://blast.ncbi.nlm.nih.gov/Blast.cgi); presence of reported escape mutations; overlap with previously reported immunogenic epitopes (D-Overlap) retrieved using an internal database. (*Multiple HLA affinity*) following these filtration layers, Custommune identifies whether any predicted epitope displays high-affinity to multiple HLA alleles and (*Final epitope filtration*) discards any epitopes that have reported escape mutations and/or are not located in an evolutionary conserved region. (*Affinity robustness*) among remaining candidates, Custommune restricts further analyses on the three top scoring epitopes for both HLA classes. For these, Custommune computes the HLA binding affinities of potential mutant versions, though not classified as escape mutations, to estimate the impact of these mutations on epitope recognition (SDaffinities). (*HLA-epitope docking*) on the same three top ranking epitopes, Custommune computes epitope-HLA allele docking scores, calculated using the LightDock^79^ python package and scored using the DFIRE^85^ scoring function. (*Final output and annotation*) in a parallel process, the Bepipred 2.0^39^ algorithm is implemented to predict neutralizing antibody epitopes from the initial consensus sequence, that can be further intersected with Class II restricted epitopes to increase immunogenicity. As a final output, for both Class I and II HLAs, Custommune ranks the top 3 epitopes according to a score (CustoScore) which accounts for all aforementioned filtration parameters.

For HIV-1 therapeutic vaccine design, Custommune crosses input data from patient-specific viral sequences (DNA in FASTA format or raw DNA sequencing inputs) and patient’s HLA-I and/or HLA-II alleles, providing an output of epitopes of desired *k*-mer length. To facilitate the allele input step, the tool provides two links directing the user to a list of supported Class I and Class II HLA alleles, respectively. These lists mirror those of the netMHCpan 4.0 algorithm^35^, for either HLA class. Although the approach could potentially be extended to encompass entire HIV-1 sequences, we decided to limit the search for viable epitopes to the *gag* gene only, because of the previously described distinctive efficacy of anti-Gag cell-mediated immunity^18,24–27^.

The tool pipeline (Figure 1) starts by translating input *gag* genomic sequences to protein sequences. Custommune then performs multiple sequence alignment using the Clustal Omega (REST) web service Python client^36^ and builds a consensus translated sequence. The consensus sequence is then used to predict epitopes restricted to patient-specific HLA-alleles for both classes. The HLA-specific epitopes provided as final output by Custommune are pre-filtered by the algorithm. This pre-filtering follows a set of parameters that compute epitope affinity in terms of sequence variation and conservation degree, allele-restricted affinities, and previous clinical evidence of immune response (Figure 1). For calculating evolutionary conservation, each epitope is compared, in terms of similarity, to an internal database of Gag amino acid sequences (Supplementary File 1) collected mainly from curated alignments retrieved from the Los Alamos HIV sequence database (http://www.hiv.lanl.gov/). Moreover, to verify whether antigenicity has already been reported for the candidate epitopes, the tool compares potential epitopes to those already described in the Los Alamos HIV immunology site (http://www.hiv.lanl.gov/content/immunology).

To further refine the structural assessment of epitope binding to HLA-alleles, Custommune performs structural epitope modelling followed by epitope-HLA docking to determine the structural stability of the HLA-predicted epitope binding (Figure 1). The Custommune pipeline also computes some related physicochemical parameters of the personalized epitope sequence to aid in the assessment of the structural stability of candidate peptides.

Overall, the tool is optimized to identify immunogenic peptides characterized by the lowest variability (mutation potential). In line with this, the tool specifically highlights potential epitopes that are contained in regions which were previously described as essential for viral fitness^29,37^. This is a novel and fundamental feature of this approach, as RNA viruses are characterized by a high ability to mutate^38^.

The Custommune pipeline can be applied to other vaccine strategies by following a parallel workflow (Figure 1). An example of these applications are acute infections, such as COVID-19. In this case, an approach combining neutralizing antibody responses and recognition by HLA haplotypes most represented in a given population might provide a reasonable compromise between specificity and scalability. To this aim, using Bepipred-2.0^39^, Custommune can identify potential neutralizing epitopes which overlap with epitopes consistent with recognition by population-specific Class II HLA haplotypes. At the same time, Custommune can predict another set of epitopes optimized for recognition by HLA Class I haplotypes of the same population, thus providing two levels of potential immune recognition.

Overall, the Custommune pipeline provides a flexible and fast tool to generate epitope predictions according to the genetic diversity of the virus and the genetic HLA profile/s of the host or susceptible populations.

### Correlation between Custommune predictions and therapeutic vaccine efficacy in PLWHA

We tested Custommune predictions against manual epitope selection using results from an ongoing multi-interventional phase II clinical trial enrolling PLWHA (NCT02961829)^32^. In this trial, autologous dendritic cells were pulsed with a personalized vaccine designed manually from Gag sequences generated from each patient’s circulating virus. In the study groups (G5 and G6) that had received this vaccine (along with other interventions), the patients showed variable responses including two individuals who displayed significant control of viral load during ATI^32^. When viral and HLA sequences of patients from G5 and G6 were used as input for Custommune, the epitopes predicted by the tool generally displayed some overlap with those administered in the study (Figure 2A).

**Figure 2.**
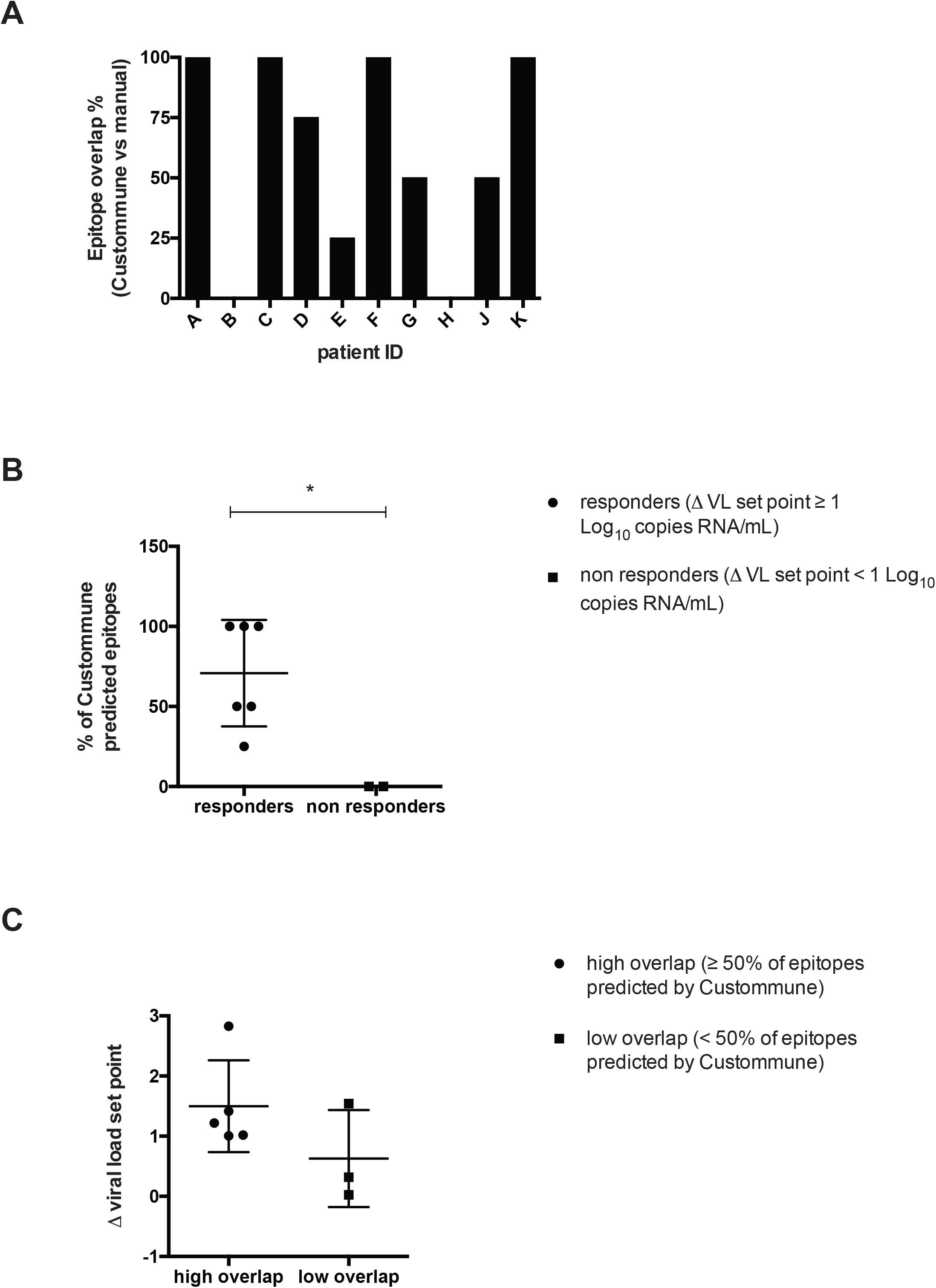
Potential therapeutic efficacy of Custommune-predicted vaccine candidates. (A) Percentage of personalized peptides predicted by Custommune which overlap with those administered as vaccines to people living with HIV/AIDS (PLWHA) in clinical trial NCT02961829. Each letter indicates a trial participant. (B) Percentage of overlap between epitopes predicted by Custommune and epitopes administered in the trial in virologic responders and non responders. Virologic responders were defined as individuals with Δ viral load set point ≥ 1 Log_10_ copies of HIV-1 RNA/mL of plasma. Data were analyzed by two-tailed Student *t*-test. Panel C) Δ viral load set point in trial participants who received peptides with high or low overlap to Custommune predictions (≥ 50% or < 50% overlap, respectively). The Δ viral load set point was calculated as the difference between pre- and post-therapy viral load set points, with post-therapy viral load set point calculated as the median of all available measurements (up to 9 weeks post-treatment interruption). Each data point in panels B and C indicates a trial participant.

Therefore, to investigate the potential therapeutic efficacy of Custommune predictions, we stratified patients based on the virologic response during ATI, which was defined as > 1 Log_10_ Δ `viral load set point` (*i*.*e*. the difference between median pre- and post-therapy copies of HIV-1 RNA/mL of plasma). Of note, non-responders were the only patients for whom there was no overlapping prediction between epitopes calculated by Custommune and those administered *in vivo* (Figure 2B). Conversely, patients who had been administered vaccine epitopes highly overlapping (>50%) with those predicted by Custommune, were characterized by higher viral load abatement (Figure 2C). These data suggest that Custommune can predict epitopes with therapeutic potential and could improve both efficacy and speed of personalized vaccine design.

### Identification of input SARS-CoV-2 sequences for Custommune

As the ongoing COVID-19 outbreak is an urgent challenge for vaccine development^40^, we decided to test the potential of Custommune for rapid identification of vaccine targets. In order to utilize Custommune for SARS-CoV-2 predictions we first decided to identify the viral regions that could act as optimal input for the tool.

Due to the recent evolution of SARS-CoV-2, there is no equivalent of HIV-1 Gag, *i*.*e*. a validated viral target for effective immunity. However, SARS-CoV-2 shares approximately 80% sequence identity with SARS-CoV^41^, the causative agent of an epidemic burst of acute respiratory distress syndrome (ARDS) in 2003. Therefore, we decided to use previously described strategies successfully targeting SARS-CoV replication as a template to restrict Custommune predictions. In particular, our efforts were directed at two validated sub-targets within the S-glycoprotein necessary for viral attachment to host cells^42^: 1) the portion of the S-glycoprotein that mediates the main protein-protein interaction with the cellular entry receptor, *i*.*e*. angiotensin converting enzyme 2 (ACE2), as this was described as an optimal target for neutralizing antibodies^43^; 2) the viral S-glycoprotein region binding the glycosylated portion of ACE2, an interaction inhibited by pretreatment with chloroquine^44,45^, a drug recently shown to effectively hamper SARS-CoV-2 replication *in vitro* and in patients^46,47^.

In order to translate these approaches into vaccine design:

1. We performed a thorough analysis for molecular complexes of the viral S-glycoprotein with the entry receptor ACE2. Considering the configuration of ACE2, we superimposed complexes of S-glycoprotein/ACE2 in both states of the receptor, *i*.*e*. free or bound (in this case with the competitive inhibitor MLN-4760)^48^. Our analyses indicated that the receptor-binding domain (RBD) surface of S-glycoprotein interacting with the bound configuration of ACE2 is relatively smaller than (though 100% overlapping with) that interacting with the unbound configuration of ACE2 (Figure 3A,B). In light of this, we decided to restrict the Custommune input to the RBD sequence interacting with bound ACE2 and the linker amino acids (henceforth RBDp) (Figure 3A,B). It is expected that this approach will be able to evoke antibodies against the RBDp irrespective of the ACE2 bound/unbound configuration.
2. We inspected the possible contribution of oligosaccharide moieties of ACE2 to the S-glycoprotein/ACE2 binding interface. The oligosaccharide moiety of ACE2 was described as fundamental for optimal binding of the S-glycoprotein of SARS-CoVs^44^. So far, in published structures, only partial ACE2-bound oligosaccharide data is available. Therefore, we decided to study this phenomenon by analyzing a published structure of inhibitor-bound ACE2 (1R4L), which presents an N-acetylglucosamine (NAG) covalently bound to residue Asn90 and remaining from the oligosaccharide originally attached to this protein^49^. This evidence suggests that the NAG present in the 1R4L structure is a marker of the position of the oligosaccharide originally attached to ACE2 before being altered by the crystallization process. By superimposing this structure to the structure of the S-glycoprotein with ACE2 and measuring the atomic distances at the binding interface between NAG and the S-glycoprotein, we were able to determine the specific segment of the S-glycoprotein RBD that could be responsible for the interaction with the ACE2-bound oligosaccharide. Two specific residues of the S-glycoprotein (Gly416 -Lys417) were found to interact with NAG, being within a 10 Å radius from NAG, *i*.*e*. a distance associated with significant intermolecular interactions (Figure 3C,D). Using S-glycoprotein Gly416 as a starting point, we selected a core peptide spanning 20 amino acids in both directions of the translation frame. This led to the identification of a segment of the S-glycoprotein RBD, which we henceforth name RBDg, as a *bona fide* target for vaccine epitope design (Figure 4A).

Of note, a structure of the SARS-CoV-2 S-glycoprotein and ACE2 interaction (PDB: 6M17) was recently published while the present report was in preparation^50^. The authors concluded that the binding interface to ACE2 is similar for SARS-CoV and SARS-CoV2, and their conclusions are largely overlapping with the results of the present analyses.

**Figure 3.**
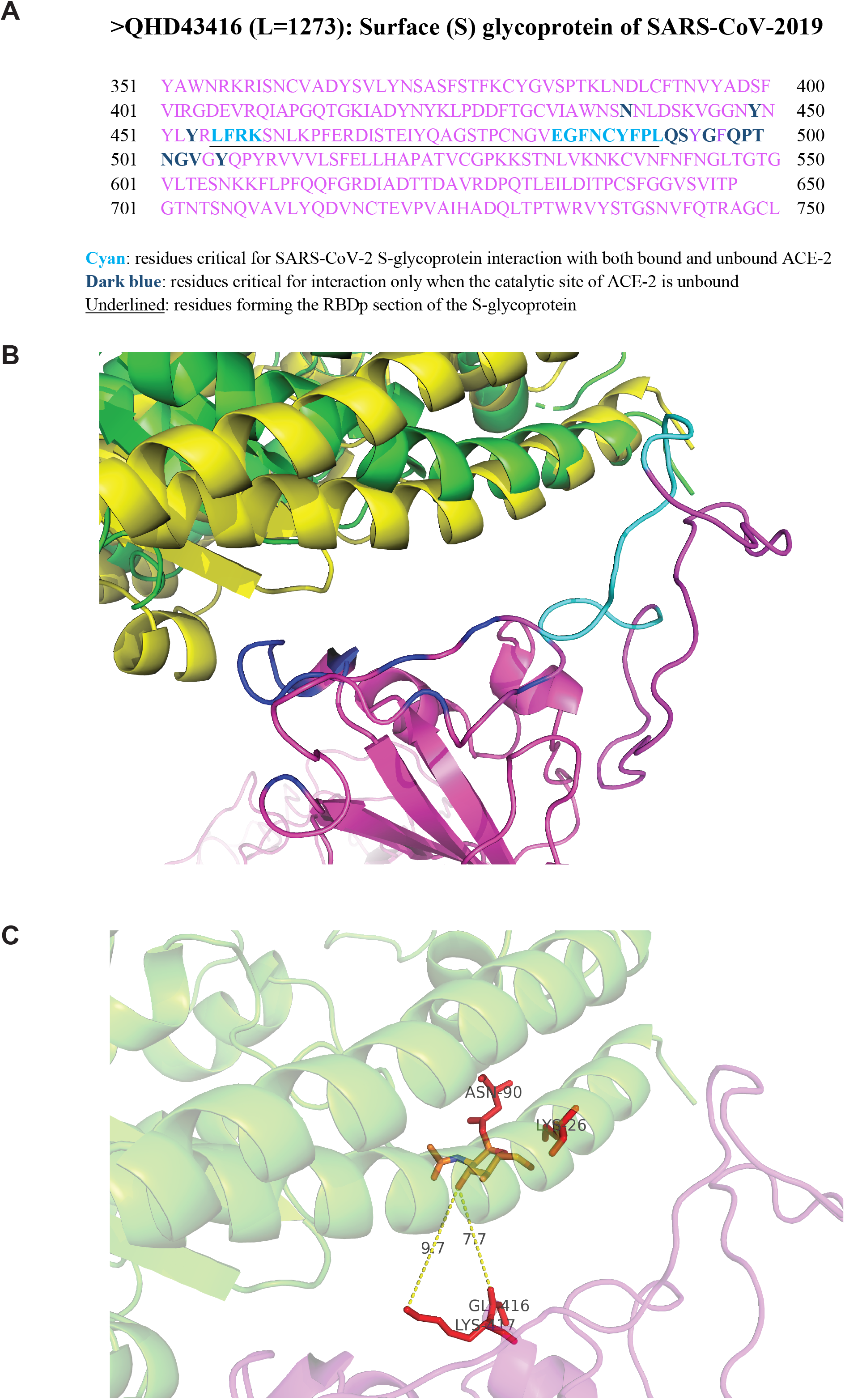
Identification of vaccine targets in the receptor binding domain (RBD) of the SARS-CoV-2 Spike (S) glycoprotein. (A) Partial sequence of the SARS-CoV-2 S-glycoprotein (derived from structure QHD43416^90^). Residues constituting the protein-protein interaction surface of the S-glycoprotein (magenta) with ACE2 are shown in different gradations of blue. Residues responsible for binding of the S-glycoprotein only in the presence of unbound catalytic site of ACE2 are shown in dark blue. The residues underlined correspond to the receptor binding domain 1 (RBDp), as described in the main text. (B) Interaction of SARS-CoV-2 S-glycoprotein (magenta) with superimposed structures of unbound ACE2 (yellow) or ACE-2 bound to the competitive inhibitor MLN-4760 (green). The specific segment in the receptor binding domain (RBD) of the S-glycoprotein that was found to overlap with both configurations of ACE2, *i*.*e*. unbound catalytic domain or catalytic domain bound with inhibitor MLN-4760, is shown in cyan. Residues binding only to unbound ACE-2 are shown in dark blue. (C) Proximity of N-acetyl-D-glucosamine (NAG) (shown in CPK) to the interaction interface between the spike glycoprotein and ACE2. Asn90-bound NAG in ACE2 was found to interact with Lys26 of ACE2 and Gly416 and Lys417 of the S-glycoprotein.

**Figure 4.**
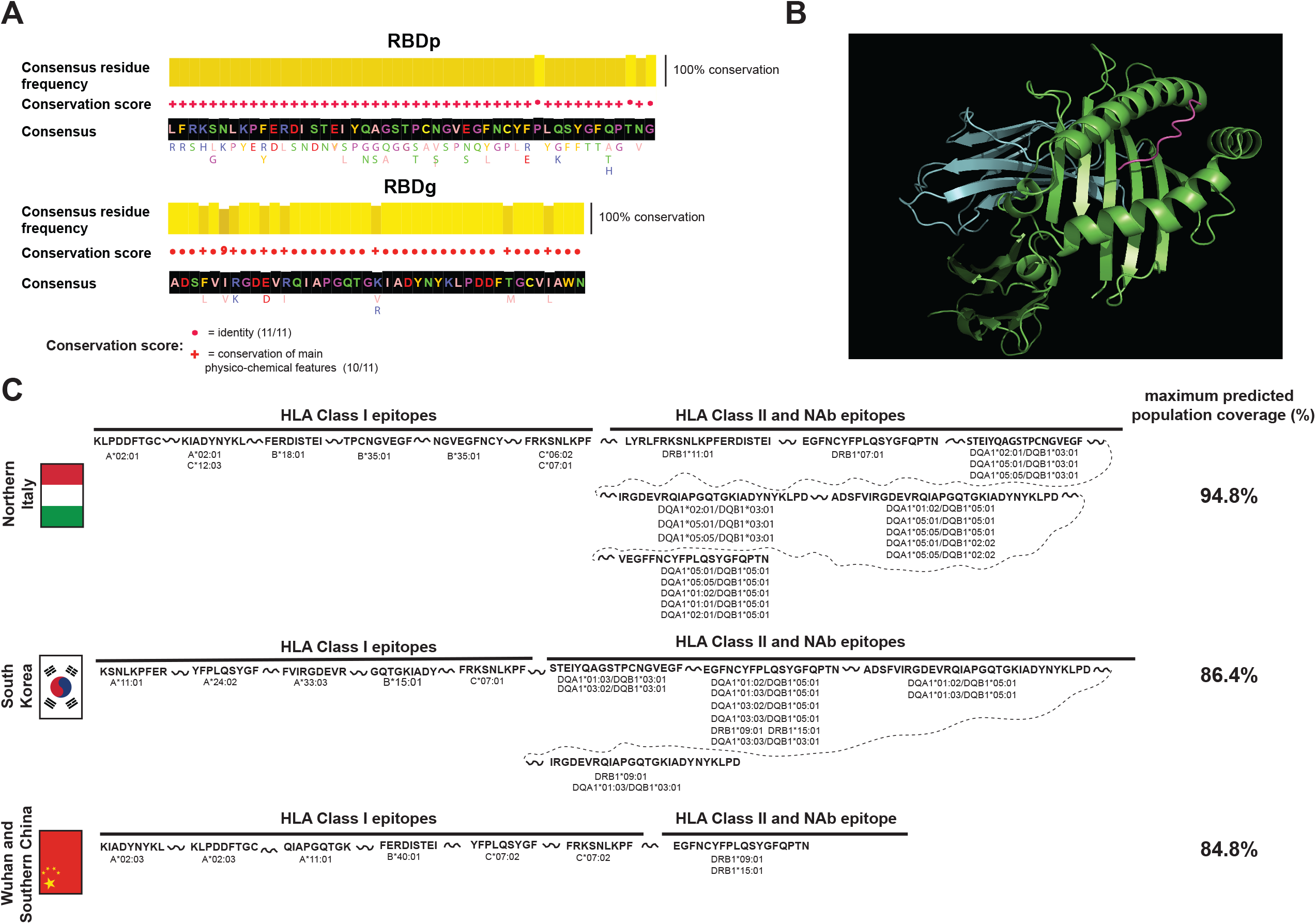
Population-targeted vaccine design against the RBDp and RBDg regions of SARS-CoV-2. (A) Evolutionary conservation of RBDp and RBDg regions of the S-glycoprotein of SARS-CoV-2. Consensus sequence and evolutionary conservation were calculated based on the multiple sequence alignments in Supplementary Files 2 and 3 using Jalview^87^. The conservation score is based on^51^. (B) Example of epitope-HLA docking pose generated using LightDock^79^. The Custommune-predicted epitope “KIADYNYKL” (magenta) is shown restricted by the HLA class I histocompatibility antigen A-2 α-chain (HLA-A*02:01, green), which is highly expressed in Northern Italy (Supplementary File 4). Also shown is the invariant β_2_-microglobulin (cyan). The docking pose was scored using the DFIRE function^85^ as listed in Supplementary file 5. (C) Custommune vaccine predictions and expected coverage for each target population. Predicted epitopes were selected from those on which docking was performed (Supplementary File 5). Maximum expected population coverage was calculated based on allele frequencies in each population (listed in Supplementary File 4) according to the formula described in the “Materials and Methods” section. Linker regions between vaccine peptides are an example of a vaccine strategy based on a single, multi-epitope, formulation.

Overall, this evidence shows that the RBDp and RBDg DNA sequences of SARS-CoV-2 can be used as optimal inputs for Custommune.

### Custommune epitope predictions for population-targeted SARS-CoV-2 vaccines

To mimic the approach described for HIV-1, we first analyzed the variability of RBDp and RBDg by multiple alignment of all SARS-CoV-2 S-glycoprotein sequences available at NCBI and GISAID (including isolates from humans, bats and pangolins) (Supplementary File 2, 3 and Figure 4A). In line with the predicted key structural role of RBDp and RBDg, both sequences displayed very limited variability, mostly deriving from non-human isolates (Supplementary File 2 and 3). Moreover, every amino acid variant (except one in RBDg) fully preserved the main physico-chemical characteristics of the consensus residue (according to the scoring system of ^51^). These results suggest that both RBDp and RBDg represent *bona fide* equivalents of the conserved Gag sequences used as privileged targets for Custommune HIV-1 predictions.

To adapt Custommune predictions to some of the populations most affected by the SARS-CoV-2 pandemic (at the time at which these analyses were performed), we retrieved the relative HLA allele frequencies in individuals from Northern Italy and South Korea (Supplementary File 4) (Allele Frequency Net Database; http://www.allelefrequencies.net)^52^. Moreover, we applied the same approach to HLA alleles of individuals from Southern China and from the city of Wuhan, where the outbreak had initially spread (Supplementary File 4).

When the RBDp and RBDg sequences were used as inputs along with population-specific HLAs, Custommune returned a set of epitopes (Supplementary File 5) for either Class I or Class II HLAs. The HLA Class II specific epitopes were further filtered to highlight those predicted as targets for neutralizing antibodies using Bepipred-2.0^39^. This was done to ensure that a unique peptide may provide the double stimulus necessary for optimal B-cell activation and antibody production (Supplementary File 5).

In line with the Custommune pipeline, and in order to improve the likelihood of immune recognition, the binding stability and affinity of the most promising epitopes was validated by molecular docking. In particular, epitopes were selected for docking if they had been predicted to bind with an IC50 < 600 nM^53^ to HLA alleles described at four digit resolution for the population of interest in the Allele Frequency Net Database (Supplementary File 5 and Figure 4B,C). Interestingly, the identified epitopes included key residues involved in hydrogen bond formation between the S-glycoprotein of SARS-CoV-2 and ACE2 (*e*.*g*. Gln 474 in epitope STEIY<u>Q</u>AGSTPCNGVEG, Gln498 in epitope LQSYGF<u>Q</u>P and Lys417 in epitope IRGDEVRQIAPGQTG<u>K</u>IADYNYKLPD of S-glycoprotein, engaged, respectively, in hydrogen bonds with residues Gln24, Tyr41, Asp30 of ACE2). Since hydrogen bonds were recently described as crucial for the stability of the virus-receptor interaction^50^, epitopes containing the hydrogen-bonding residues might be particularly suitable targets to evoke immunity against structural determinants of SARS-CoV-2 infection. Moreover, in order to ensure the best coverage likelihood of the target population, we also included the predicted epitopes for the most prevalent Class I HLA antigens. Our results show that a peptide set specific for both neutralizing antibody/HLA Class II and for HLA Class I could provide a good population coverage upon simultaneous delivery, potentially achieving herd immunity (Fig 4C). Of note, one of the most promising epitope candidates designed by Custommune for two of the populations examined (*i*.*e*. epitope KLPDDFTGC for Southern China/Wuhan and Northern Italy) (Supplementary File 5 and Figure 4C) is equivalent to a highly immunogenic peptide previously identified by stimulating cells of patients who had successfully recovered from SARS infection^54^.

Taken as a whole, these results show the application of Custommune to predict epitopes for specific populations and highlight a set of vaccine candidates to curb the spread of SARS-CoV-2 in highly affected areas.

## Discussion

The precision medicine era, albeit still in its early stages, is expected to supersede traditional, one-size-fits-all therapeutic approaches. The development of personalized, yet scalable, treatments would allow accounting for the genetic variability of individuals, pathogens, or cancer profiles, and pave the way for more accurate efficacy predictions while reducing side effects. The implementation of our Custommune pipeline in the context of HIV/AIDS shows that the tool algorithm may be used to predict novel immune-based treatments with *in-vivo* potential. Even though the pipeline was applied to the HIV-1 Gag protein in the present work, it can potentially be extended to other HIV genomic regions or other chronic viral infections. Crucial pre-requirements of the personalized Custommune approach are the identification of a key structural component of the target pathogen and the obtainment of sequencing data from both the host HLA alleles and the infecting virus. While cost considerations might represent a limiting factor in some settings, the quick advances in sequencing technology, coupled to the steep reduction in price^55^, make the approach already feasible in developed countries. Moreover, a personalized intervention aimed at a cure could make the cost-benefit analysis attractive also in developing countries, which often bear the main burden of chronic viral infections^56^.

In terms of potential efficacy, the Custommune approach relies on minimizing epitope diversity while maximizing predicted binding strength and immunogenicity of said epitopes. It is noteworthy that, when compared to a real clinical scenario, epitopes predicted by Custommune correlated with treatment response. This was likely aided by the large amount of immunologic data available on HIV-1 (*e*.*g*. Los Alamos HIV immunology site). Therefore, due to its low cost and scalability, Custommune could be immediately applied to the design of therapeutic HIV peptide vaccines^57^ or autologous dendritic cell vaccines pulsed with tailored Gag peptides. Compared to previous attempts at streamlining vaccine design in the context of cancer^58^, the Custommune pipeline includes multiple layers of epitope ranking with scoring parameters accounting for: mutational potential, structural conservation, HLA docking, escape mutations, location of the neo-mutation and previous evidence of antigenicity. These partially redundant filtration stages are envisaged to maximize the chances for durable and potent epitope recognition. Moreover, other filters such as predicted epitope processing and cleavage have been included to the pipeline when this manuscript was in preparation, confirming the versatility of the Custommune approach.

Our implementation of Custommune was here extended to include vaccine design for SARS-CoV-2. Current predictions suggest that traditional vaccine strategies might be too slow to address the spread of the pandemic and mitigate the death toll^59^. Furthermore, immune responses developed during natural infection might be insufficient to provide long-term protection against reinfection^60^. The approach herein proposed is aimed at a flexible response customized for the populations most affected at a given time. As a novel pathogen will necessarily lack the wealth of immunologic data available for heavily studied viruses like HIV-1, our vaccine strategy attempts both induction of cell-mediated immunity and neutralizing antibodies. Indeed, early evidence indicates that a broad immune response might correlate with successful clearance of the infection^61^. Custommune predicted epitopes would further combine this broad immune stimulation with a design based on the most common HLA alleles in the population of interest, potentially providing enough immune coverage for the induction of herd immunity. Moreover, the choice of a highly conserved viral target as a source of vaccine epitopes should ensure a broadly effective response in those individuals for whom the vaccine should prove immunogenic. By utilizing Custommune, the whole vaccine design process should last less than a working day. Therefore, this approach, if successful, could be quickly adopted to blunt the pandemics during its spread or, ideally to pre-empt it.

The binding affinities predicted by Custommune for epitopes derived from RBDp and RBDg were generally higher for HLA Class I alleles in the populations here considered. While this is not sufficient to predict that cell mediated immunity would be preferentially induced by the proposed vaccine, previous evidence in mice suggests that memory CD8^+^ T-cells might alone be sufficient to provide effective protection against SARS-CoV^62^. Corroborating this hypothesis, one of the peptides designed by Custommune was equivalent to an epitope associated with clearance of SARS-CoV infection during the previous epidemics and with immunogenicity in mice when used as a vaccine^54^.

Our estimate of the probability to reach herd immunity in the populations considered is based on the assumption that the development of immune responses against each of the vaccine peptides would be *per se* sufficient to guarantee some level of protection. While this prerequisite might prove optimistic, it is noteworthy that the viral targets selected for vaccine design (*i*.*e*. RBDp and RBDg of the S-glycoprotein) display exceptional evolutionary conservation and that no polymorphism in these regions was detected in the viral isolates from either Italy, South Korea or China. This conservation, coupled with the generally moderate mutation rate of SARS-CoV^63^ and SARS-CoV-2^64^ as compared to other RNA viruses, yields credibility to the idea of achieving protection by targeting single immunodominant epitopes^62^. Moreover, the expected population coverage of each of the vaccines designed in the present study is theoretically sufficient to achieve herd immunity based on the estimated reproductive number of SARS-CoV-2^34,65^.

In the current work, to simplify administration schedule and increase scalability, we envisage, among other possibilities, a strategy synthesizing one multi-epitope peptide for each target population. This peptide would link Class II HLA-restricted and neutralizing antibody epitopes as well as Class I HLA-restricted CD8^+^ T-cell epitopes. However, this approach will require empirical validation and could be modified, *e*.*g*. by administering HLA Class I and Class II restricted epitopes in separate formulations. While reduced immunogenicity is a well-known caveat of epitope-based vaccines, recent advances in adjuvant and delivery technology might allow overcoming this limitation^66^. Apart from classical adjuvants, the use of an “adjuvant” drug such as chloroquine, is of particular interest for SARS-CoV-2. This treatment option could enhance vaccine immunogenicity^67,68^ while possibly providing *per se* some protection against the virus^69^. In terms of delivery, carriers such as liposomes and nanoparticles, or strategies employing chemical conjugation or cell-penetrating peptides could increase epitope presentation by antigen presenting cells^66,70^. Finally, in our current model, we envisaged the use of linker sequences with protease cleavage sites between different epitopes^71^. This strategy might increase the chances of presenting peptides of optimal size to both HLA alleles of Class I and Class II. However, covalent linkage of epitopes has also been described to increase immunogenicity^66^. *In-vivo* studies will be required to optimize these strategies for inducing immunity against SARS-CoV-2. Due to the ongoing rapid expansion of the epidemics and the relatively good safety profile of peptide vaccines^66^, pilot clinical testing in significantly affected areas might be envisaged.

Overall, our study describes a novel tool to improve multi-epitope vaccine design specificity while drastically reducing the associated time and cost. The pipeline herein described can be directly applied for testing personalized therapeutic vaccines for HIV-1 and to identify the core epitopes of preventive vaccines aimed at populations heavily affected by SARS-CoV-2.

## Materials and Methods

### Custommune design and pipeline implementation

#### a) Web application

The web application of Custommune is available at http://www.custommune.com. Written in Python (v3.7) using Django (v2.2.6) Custommune is a tool that provides an integrated pipeline (Figure 1) for prediction and filtration of personalized epitopes.

#### b) Sequence processing

The Biopython package^72^ is used for translating input sequences. Alignment of translated sequences is then performed using the Python client of Clustal Omega (REST) web service^36^. A consensus of the aligned sequences is generated using the Biopython module with a 50% similarity cutoff. The Biopython “ProteinAnalysis” function is used to estimate physicochemical parameters and secondary structure of the consensus sequence, including: molecular weight, gravity, specific count of amino acids, isoelectric point and fractions of secondary structures.

#### c) Epitope prediction and filtration layers

Custommune is connected with RESTful interface (IEDB-API)^73^ which serves as a platform for using NetMHCpan v4.0^35^ for Class I and II HLA predictions as well as Bepipred v2.0^39^ for antibody epitope predictions. The Pandas package (McKinney et al. 2010) is then used to structure epitope sorting tables and allow for comparative filtration. The primary filtration is based on IC50 values, a cutoff of 1000 nM is used to prevent loss of potentially false negatives.

The Los Alamos HIV database (http://www.hiv.lanl.gov/content/immunology) was used to create internal HLA class-specific datasets of previously reported immunogenic epitopes against HIV Gag. Using Pandas^74^, high-affinity epitopes are compared to these datasets to highlight epitopes with previously described immunogenicity. Moreover, another filtration layer is designed to report escape variants by comparing each epitope to an internal database collected from various literature sources including: dataset of HLA-associated polymorphisms in HIV-1 Gag as reported in Ref.^75^, as well as the datasets reported in Ref.^76^ and the datasets of CTL/CD8^+^ and T Helper/CD4^+^ epitope variants and escape mutations reported in the Los Alamos HIV database (http://www.hiv.lanl.gov/content/immunology/). Additional filtration is obtained by comparing the epitope location within the Gag sequence, to Gag regions essential for viral assembly and packaging, which tend to be structurally and evolutionarily conserved, as reported in Ref.^29^. To further refine this filtration, Custommune computes the degree of conservation for each epitope by comparing the epitope sequence to the HIV Sequence Compendium database^77^ which includes 680 alignments of HIV-1/SIVcpz Gag protein sequences. The degree of conservation (Cscore) of each epitope is calculated as a fraction represented by the subset of sequences{*s*}in which the epitope scored a local alignment of more than 80% using Clustal Omega^36^ over the total sequences *Stotal* in the internal database.

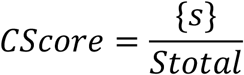

The next layer of filtration selects only epitopes that rank high for multiple alleles in case a multiple-allele input was selected by the user for both HLA classes. For further assessment of the impact of predictable mutations, Custommune computes the effect of these mutations (retrieved from the internal Gag sequence database; Supplementary File 1) on the binding affinity of epitopes to the patient HLAs. This refined analysis is performed only on the top three ranking epitopes initially predicted by the tool. By computing affinities to the same allele the user can estimate the impact of mutations in this specific segment on the affinity to the restricted allele. The degree of deviation of the mutated version is estimated based on *SDaffinities*, which are calculated as a standard deviation (SD) of the set of IC50 values for the candidate epitope and its mutant versions. The deviation value is therefore considered to negatively reflect the binding stability of this peptide segment to a restricted allele, in respect to a set of predicted mutant versions of the same segment.

#### d) Structural validation and epitope reporting

The Python package PeptideBuilder^78^ is used for generation of 3D models of top epitopes, while the package LightDock^79,80^ is implemented to perform epitope-HLA docking based on the Glowworm Swarm Optimization (GSO) algorithm^81^. Solved structures of HLA alleles were collected from the pHLA3D database^82^ and The Protein Data Bank (PDB)^83^. Homology modelling of structurally unsolved HLA alleles was generated using SWISSMODEL^84^. Distance-scaled, finite ideal-gas reference (DFIRE) function^85^ is used to calculate mean force potential of all atoms in a residue-specific manner within a resolution of less than 2 Å, which has been found to accurately predict stabilities of structural (HLA-epitope) complexes. DFIRE was implemented as a scoring function for LightDock simulations and docking scores were added in the final filtration layer for the highest ranking epitope candidates.

#### d) Final scoring and annotation

For highly ranking epitope candidates, a scoring function is designed to account for each filtration layer. In this function each continuous parameter (*IC50, DFIRE, CScore and SDaffinities*) is represented by a quantitative value, according to the following rules: 1) the IC50 value is rescaled by calculating its reciprocal multiplied by a weighting factor of 10^4^; 2) docking scores are preceded by a negative sign to weight the negative binding energies of the DFIRE scoring function of LightDock; 3) CScore is considered as a percentile of the Cscore fraction weighted by a factor of 10^3^; 4) SDaffinities are preceded by a negative sign to weight the positive values of deviation values. Categorical parameters (*LocationsScore* and *DOverlap*) are represented by binary values weighted by a factor of 500 for favorable states while non favorable states are given null values. Overall the formula to calculate the final ranking (S) can be calculated as follows:

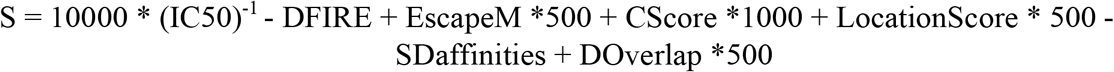

The top three epitopes ranked by S score are further analyzed based on their possible overlap with epitope data sets previously associated with: post-ART control, efficacy in vaccine studies and the lack of reported escape mutations. Finally, predicted antibody epitopes estimated by Bepipred 2.0^39^ are reported if they overlap with the top candidate epitopes ranked by S score. To allow manual inspection of results, sequence processing data and unfiltered predictions are provided in a separate section of the results page with a downloading link for a text file.

### Multiple sequence alignment and analysis

S-glycoprotein sequences were retrieved from NCBI (https://www.ncbi.nlm.nih.gov/genbank/sars-cov-2-seqs/) and GISAID^86^. Multiple alignments were performed using Clustal Omega web service^36^. Consensus sequences and sequence conservation scores and histograms were generated with Jalview (v. 2.11)^87^ according to the amino acid conservation scoring criteria described in^51^.

### Population-specific HLA allele frequencies

Class I and II HLA allele frequencies were retrieved from the Allele Frequency Net Database (http://www.allelefrequencies.net/hla.asp)^52^ using the “HLA classical allele freq search” option. Population-specific datasets (shown in Supplementary File 4) were employed to identify the most represented HLA alleles in areas heavily affected by SARS-CoV-2 spread, namely Northern Italy, South Korea and China (Wuhan and Southern China). For all alleles analyzed, each data set provided values of frequency, which were determined as the number of copies of a given allele (X) divided by the total number of alleles in the population (of size N) assayed (*i*.*e*. frequency = X/2N). For each population of interest, only HLA alleles with frequency ≥ 0.1 in at least one dataset of the same population were considered for further analysis. When a given HLA allele was represented in more than one dataset of the same population, a weighted frequency was calculated. Specifically, given an allele of interest represented in n datasets with population sizes N_1_, N_2_…N_n_, with a frequency of F_1_, F_2_…F_n_, the weighted frequency (F_w_) of the allele was calculated as:

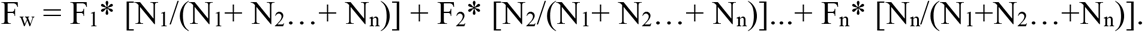

As the datasets employed included HLAs characterized at different resolutions, allele frequencies were considered separately in case a 2 or ≥4 digit resolution^88^ was available (Supplementary File 4). Alleles at 4 digit resolution and ≥ 0.1 (weighted) frequency were used as direct input for Custommune. Alleles at 2 digit resolution and ≥ 0.1 (weighted) frequency were instead analyzed with Custommune by including all potential second field^88^ options currently supported by Custommune.

### Estimation of candidate SARS-CoV-2 vaccines population coverage

Class I and Class II HLA alleles which were predicted by Custommune to bind RBDp and RBDg epitopes of SARS-CoV-2 were used to estimate potential vaccine coverage in the populations of interest. To this aim, only (weighted) frequencies of HLA alleles available at four digit resolution in the population of interest were included (Supplementary File 4). Moreover, among these alleles, only those with high predicted binding affinity (IC50 < 600 nM) for an RBDp or RBDg epitope were included in the vaccine design (Supplementary File 5). To estimate the percentage of individuals (P) of a given population expected to carry an HLA allele, the (weighted) frequency of that allele (F) in the same population (Supplementary File 4) was used, according to the formula:

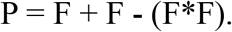

For heterodimers (*e*.*g*. HLA-DQA1 and DQB1) an overall frequency of the heterodimer was first calculated as: frequency of heterodimer 1 * frequency of heterodimer 2. This overall heterodimer frequency was then used to calculate P as described above.

Given a vaccine of N epitopes recognized by HLA alleles carried respectively by a percentage of individuals of the target population P_1_, P_2_…P_N_, the maximum theoretical population coverage (M) of the vaccine was calculated as:

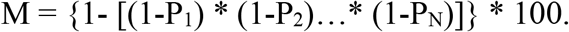

HLA alleles of the population that were predicted to recognize more than one epitope of the vaccine were considered only once in the calculation of M.

### Clinical data

HIV-1 viral loads of individuals enrolled in trial NCT02961829 were measured by q-PCR as described in^89^.

### Statistical analysis

Clinical data were analyzed by unpaired *t*-test using Graphpad Prism (v. 6 GraphPad Software, La Jolla California USA).

## Data Availability

The Custommune platform is available at http://www.custommune.com. Data used for multiple sequence alignment were retrieved from NCBI (https://www.ncbi.nlm.nih.gov/genbank/sars-cov-2-seqs/) and GISAID (https://www.gisaid.org). The structures used for homology modelling or molecular docking were retrieved from the pHLA3D database (https://www.phla3d.com.br) and the Protein Data Bank (https://www.rcsb.org). Allele frequencies were retrieved from the Allele Frequency Net Database (http://www.allelefrequencies.net/hla.asp).

https://www.ncbi.nlm.nih.gov/genbank/sars-cov-2-seqs/

https://www.gisaid.org

https://www.phla3d.com.br

https://www.rcsb.org

http://www.allelefrequencies.net/hla.asp

## Acknowledgements

MEH acknowledges support from the National Research Centre, and the Science and Technology Development Fund (STDF), Ministry of Higher Education and Scientific Research Cairo, Egypt (Grant 25632). RSD acknowledges support from the Fundação de Amparo à Pesquisa do Estado de São Paulo (FAPESP; 2013/11323-5) and the Conselho Nacional de Desenvolvimento Científico e Tecnológico (CNPq; 454700/2014-8).

The authors thank Dr. Lara Gallucci and Dr. Zunamys Carrero for critical reading of the manuscript and helpful suggestions.

## Author Contributions

Conceived the study: MT, ILS, AS; coding and web platform: MT; clinical data: JM, RD; designed Custommune: MT, MEH, ILS, AS; wrote the manuscript: MT, ILS, AS.

## Conflict of interest

MT, MEH, ILS, RD and AS have requested patent rights on Custommune and/or personalized HIV-1 vaccine design strategies.

